# Building a Best-in-Class Automated De-identification Tool for Electronic Health Records Through Ensemble Learning

**DOI:** 10.1101/2020.12.22.20248270

**Authors:** Karthik Murugadoss, Ajit Rajasekharan, Bradley Malin, Vineet Agarwal, Sairam Bade, Jeff R. Anderson, Jason L. Ross, William A. Faubion, John D. Halamka, Venky Soundararajan, Sankar Ardhanari

## Abstract

The natural language portions of electronic health records (EHRs) communicate critical information about disease and treatment progression. However, the presence of personally identifiable information (PII) in this data constrains its broad reuse. Despite continuous improvements in methods for the automated detection of PII, the presence of residual identifiers in clinical notes requires manual validation and correction. However, manual intervention is not a scalable solution for large EHR datasets. Here, we describe an automated de-identification system that employs an ensemble architecture, incorporating attention-based deep learning models and rule-based methods, supported by heuristics for detecting PII in EHR data. Upon detection of PII, the system transforms these detected identifiers into plausible, though fictional, surrogates to further obfuscate any leaked identifier. We evaluated the system with a publicly available dataset of 515 notes from the I2B2 2014 de-identification challenge and a dataset of 10,000 notes from the Mayo Clinic. In comparison with other existing tools considered best-in-class, our approach outperforms them with a recall of 0.992 and 0.994 and a precision of 0.979 and 0.967 on the I2B2 and the Mayo Clinic data, respectively. The automated de-identification system presented here can enable the generation of de-identified patient data at the scale required for modern machine learning applications to help accelerate medical discoveries.

## Introduction

The widespread adoption of electronic health records (EHRs) by healthcare systems has enabled digitization of patient health journeys. While the structured elements of EHRs (e.g., health insurance billing codes) have been relied upon to support the business of healthcare and front office applications for decades, the unstructured text (e.g., history & physical notes and pathology reports) contains far richer and nuanced information about patient care, supporting novel research^1–5^. However, this text often contains personally identifiable information (PII) as defined in the Health Insurance Portability and Accountability Act of 1996 (HIPAA), such as the personal name, phone number, or residential address^6^. As a consequence, such data has limited reuse for secondary purposes^7^. HIPAA permits data derived from EHRs to be widely shared and used when it is de-identified. Under the HIPAA Privacy Rule, de-identification can be accomplished in several ways. The most straightforward is the Safe Harbor implementation, which necessitates removal of an enumerated list of 18 categories of direct-(e.g., Social Security Number) and quasi-identifiers (e.g., date of service).

Implementing a scalable method for de-identification has several competing requirements. First, from a regulatory perspective, it must achieve extremely high recall, in that it needs to detect nearly all instances of PII. Second, from a clinical utility perspective, it must achieve extremely high precision, so that we maximize the correctness of biomedical research performed. And, third, the approach needs to be cost effective, so that millions of records can be de-identified in a reasonable amount of time. The traditional approach of manual detection of PII is expensive, time consuming and prone to human error^8,9^, which makes automated de-identification a more promising alternative^10,11^.

Several recent advances in natural language processing (NLP) have created an opportunity to build accurate and scalable automated de-identification systems. First, transfer learning of autoregressive and autoencoder models^12^ for a supervised task such as named entity recognition (NER) requires very little labelled data, reducing human effort and error. Second, attention-based deep learning models, such as transformers^13^, allow for the non-sequential processing of text and enable the generation of rich contextualized word representations. Third, semantic segmentation algorithms generate a subword-based vocabulary^14,15^ which can capture out-of-vocabulary words. Finally, the traditional transformer architecture has been improved upon through bidirectional encoder representations from transformers (BERT)^16^ and similar technologies that jointly train a *masked language model* (MLM) pre-training objective and a *next sentence prediction* task. BERT has set the stage for learning context independent representations of terms in text, and training context-sensitive models that transform those representations into context-aware representations based on the occurrence of a term in a sentence. We leverage these advances to support de-identification, which we formulate as a named entity recognition problem.

In this paper, we integrate a collection of approaches, blending the beneficial aspects of modern deep learning along with rules and heuristics, to create a best-in-class approach to automated de-identification. The system transforms each detected PII instance into a suitable surrogate to mitigate the risk that any residual PII can be used to re-identify patients **(Fig. 1)**. The nference de-identification tool can be accessed at https://academia.nferx.com/deid/.

**Fig. 1.**
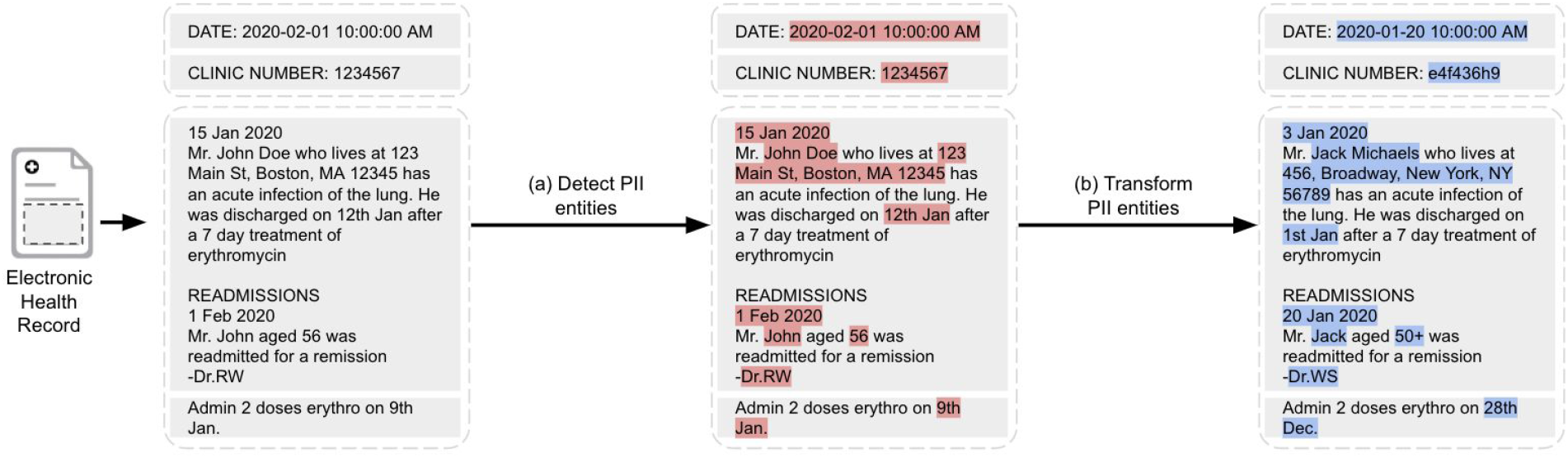
Automated de-identification of EHRs involves two steps: (a) Detecting PII entities and (b) Transforming them by replacement with suitable surrogates.

## Results

We first compare the performance of the nference de-identification system with other methods on the I2B2 2014 dataset^17^. The resulting models are evaluated using precision, recall and F1-scores (formulation provided in the Supplementary Methods) for NER on several groups of PII as defined in **Table 1**. We then compare the performance of these models on a substantially larger and diverse dataset from the Mayo Clinic and perform a deeper dive into the types of errors, distribution of errors per physician note and the distribution of errors per note type. It should be noted that this analysis focuses solely on the performance of detecting PII instances and does not address the risk of re-identification based on the semantics of any instances that the system fails to detect, an issue that is beyond the scope of this study.

**Table 1:**
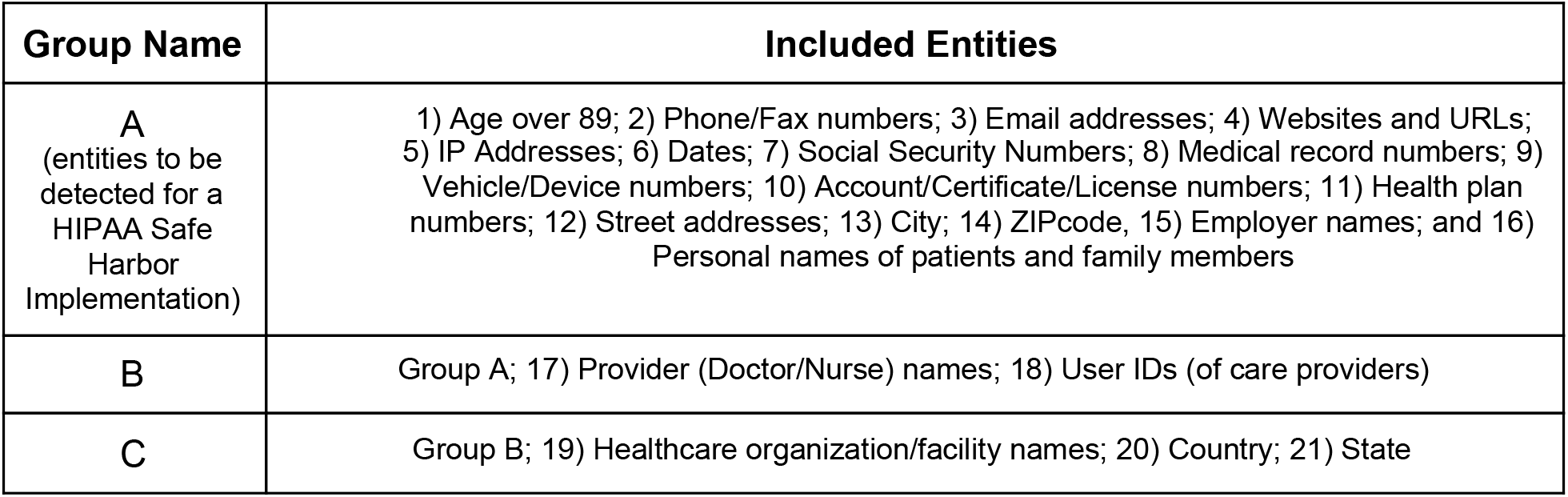
The list of entities covered by each group of direct and quasi-identifiers. It should be noted that groups B and C encompass entities beyond HIPAA Safe Harbor.

| Group Name | Included Entities |
| --- | --- |
| A<br>(entities to be detected for a HIPAA Safe Harbor Implementation) | 1) Age over 89; 2) Phone/Fax numbers; 3) Email addresses; 4) Websites and URLs; 5) IP Addresses; 6) Dates; 7) Social Security Numbers; 8) Medical record numbers; 9) Vehicle/Device numbers; 10) Account/Certificate/License numbers; 11) Health plan numbers; 12) Street addresses; 13) City; 14) ZIPcode, 15) Employer names; and 16) Personal names of patients and family members |
| B | Group A; 17) Provider (Doctor/Nurse) names; 18) User IDs (of care providers) |
| C | Group B; 19) Healthcare organization/facility names; 20) Country; 21) State |

### Performance on the 2014 I2B2 de-identification dataset

The I2B2 2014 De-identification and Heart Disease Risk Factors challenge^17^ is a publicly available dataset of clinical documents with annotated PII elements. This dataset consists of a training set of 792 clinical notes and a test set of 515 clinical notes.

We compared the performance of our approach on the 2014 I2B2 test set with six other established de-identification tools: the method proposed by Dernoncourt et al. that blends conditional random fields (CRFs) and artificial neural networks (ANNs)^18^, Scrubber^19^, Physionet^8^, Philter^20^, MIST^21^ and NeuroNER^22^

The results are provided in **Table 2**. Firstly, we cite the conditional random field and artificial neural network approach (CRF+ANN)^18^ scores against the group A entities (HIPAA only) as reported in their paper. We also directly report the results for Scrubber, Physionet, and Philter from prior publications^20^ without performing an empirical analysis because the dataset (2014 I2B2) and the set of PII entities is the same as that used in our investigation. We trained MIST using sentences from the I2B2 training corpus (see Supplementary Methods and **Supplementary Table 3**). We downloaded and used a pre-trained model for NeuroNER (see Supplementary Methods). We present the performance of these methods on group B (see **Table 1**) entities which we use as the basis of our comparison.

**Table 2:**
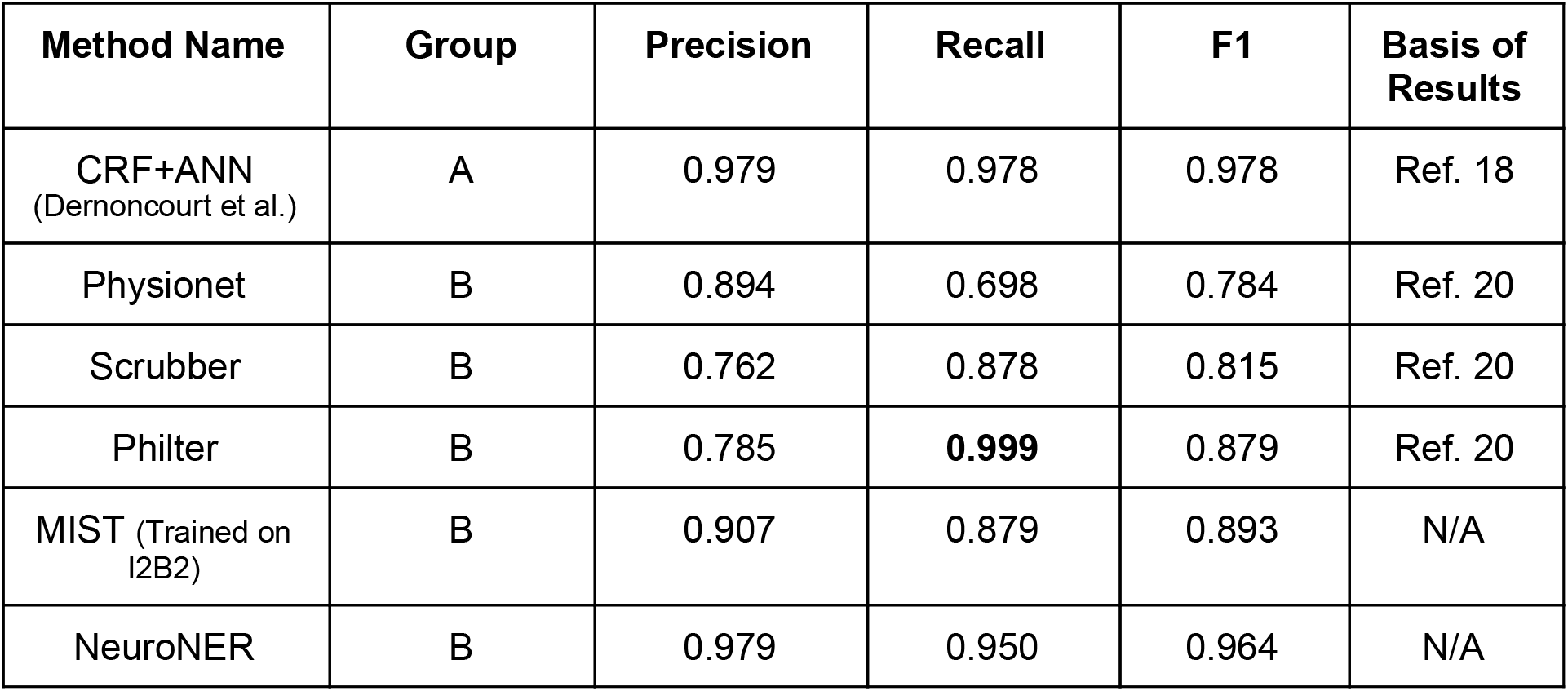
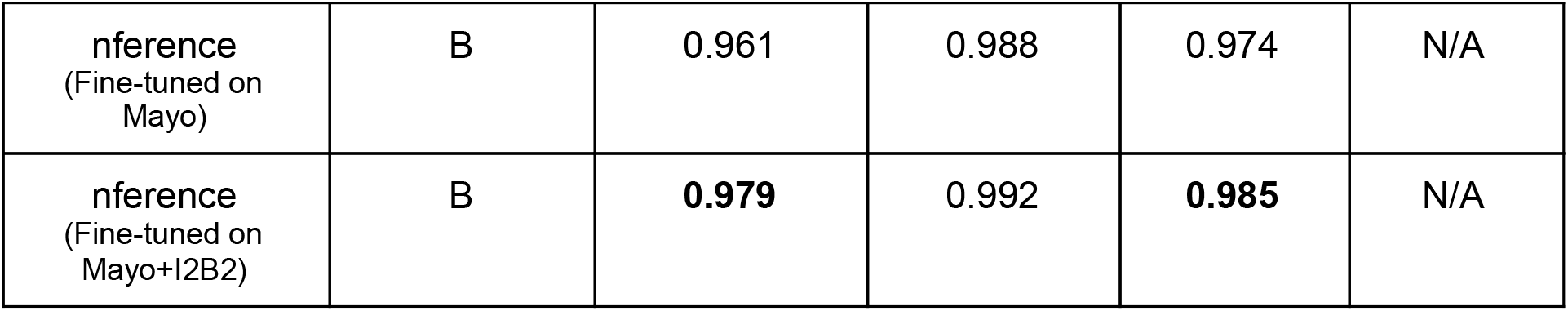
Performance of de-identification methods on the 2014 I2B2 test corpus. The results for Scrubber, Physionet, Philter and the CRF+ANN method are based on previous publications. The MIST method required training and, thus, was trained on the 2014 I2B2 training dataset. We used a pre-trained model for NeuroNER. The two versions of the nference approach were fine-tuned on (i) only the Mayo dataset and (ii) both the Mayo and I2B2 datasets.

| Method Name | Group | Precision | Recall | F1 | Basis of Results |
| --- | --- | --- | --- | --- | --- |
| CRF+ANN<br>(Dernoncourt et al.) | A | 0.979 | 0.978 | 0.978 | Ref. 18 |
| Physionet | B | 0.894 | 0.698 | 0.784 | Ref. 20 |
| Scrubber | B | 0.762 | 0.878 | 0.815 | Ref. 20 |
| Philter | B | 0.785 | **0.999** | 0.879 | Ref. 20 |
| MIST (Trained on I2B2) | B | 0.907 | 0.879 | 0.893 | N/A |
| NeuroNER | B | 0.979 | 0.950 | 0.964 | N/A |
| nference<br>(Fine-tuned on Mayo) | B | 0.961 | 0.988 | 0.974 | N/A |
| nference<br>(Fine-tuned on Mayo+I2B2) | B | **0.979** | 0.992 | **0.985** | N/A |

We present two versions of the nference system. The first version was fine-tuned only on Mayo data and did not utilize any characteristics of the I2B2 training data. When evaluated with group B, this model achieved a precision, recall, and F1 score of 0.961, 0.988, and 0.974, respectively. The second version of our system involved fine-tuning our model with sentences from the I2B2 training set. We could not incorporate inclusion lists and sentence templates associated with the I2B2 data since the dataset is small (see Methods section for details). The precision, recall, and F1 score increased to 0.979, 0.992, and 0.985, respectively. Precision and recall per identifier type is provided in **Supplementary Table 4**.

### Performance on the Mayo test dataset

The Mayo Clinic dataset consisted of 10,000 randomly sampled notes from a corpus of 104 million notes corresponding to 477,000 patients’ EHR records.

The evaluation performed on the Mayo test dataset was based on identifiers defined by group C since this group best represented the distribution of PII in the dataset. The performance of the de-identification methods (in terms of precision, recall and F1) are presented in **Table 3**. The nference method performed best with precision, recall, and F1 scores of 0.967, 0.994, and 0.979, respectively. Compared to the performance on the I2B2 dataset, we see improved recall (increase of 0.01) and a reduced precision value (decrease of 0.021). NeuroNER achieves precision, recall and F1 scores of 0.928, 0.933 and 0.931, respectively. The F1 scores of Scrubber, Physionet and Philter were lower than those achieved on the I2B2 dataset. Among these three methods, Philter demonstrates a relatively high recall of 0.918. Closely following Philter, the MIST model achieves a recall of 0.889 with overall performance similar to that on the I2B2 dataset.

**Table 3:** Precision, Recall and F1-Score of various de-identification methods on the Mayo test dataset. These methods were evaluated against group C entities.

| Method | Precision | Recall | F1 |
| --- | --- | --- | --- |
| Scrubber | 0.756 | 0.677 | 0.715 |
| Philter | 0.709 | 0.918 | 0.800 |
| Physionet | 0.837 | 0.772 | 0.803 |
| MIST<br>(Trained on Mayo) | 0.818 | 0.889 | 0.852 |
| NeuroNER<br>(Trained on Mayo) | 0.928 | 0.933 | 0.931 |
| nference<br>(Fine-tuned on Mayo) | **0.967** | **0.994** | **0.979** |

### Error analysis on the Mayo dataset

We further investigated cases in the Mayo dataset where the nference de-identification model failed to successfully detect the PII element completely (i.e., false negatives). This occurred at a rate of 0.6% (see **Table 4**). Across the 10,000 notes considered in the test set, there were 848 error instances that contained these false negative errors. Accounting for duplicate occurrences of the same sentence, there were 797 unique error instances. We grouped these instances based on the type of identifier. The prevalence of the error category is shown in the second column while the third column in the table represents the contribution of each category to the error in recall (sums to 0.6%).

**Table 4:**
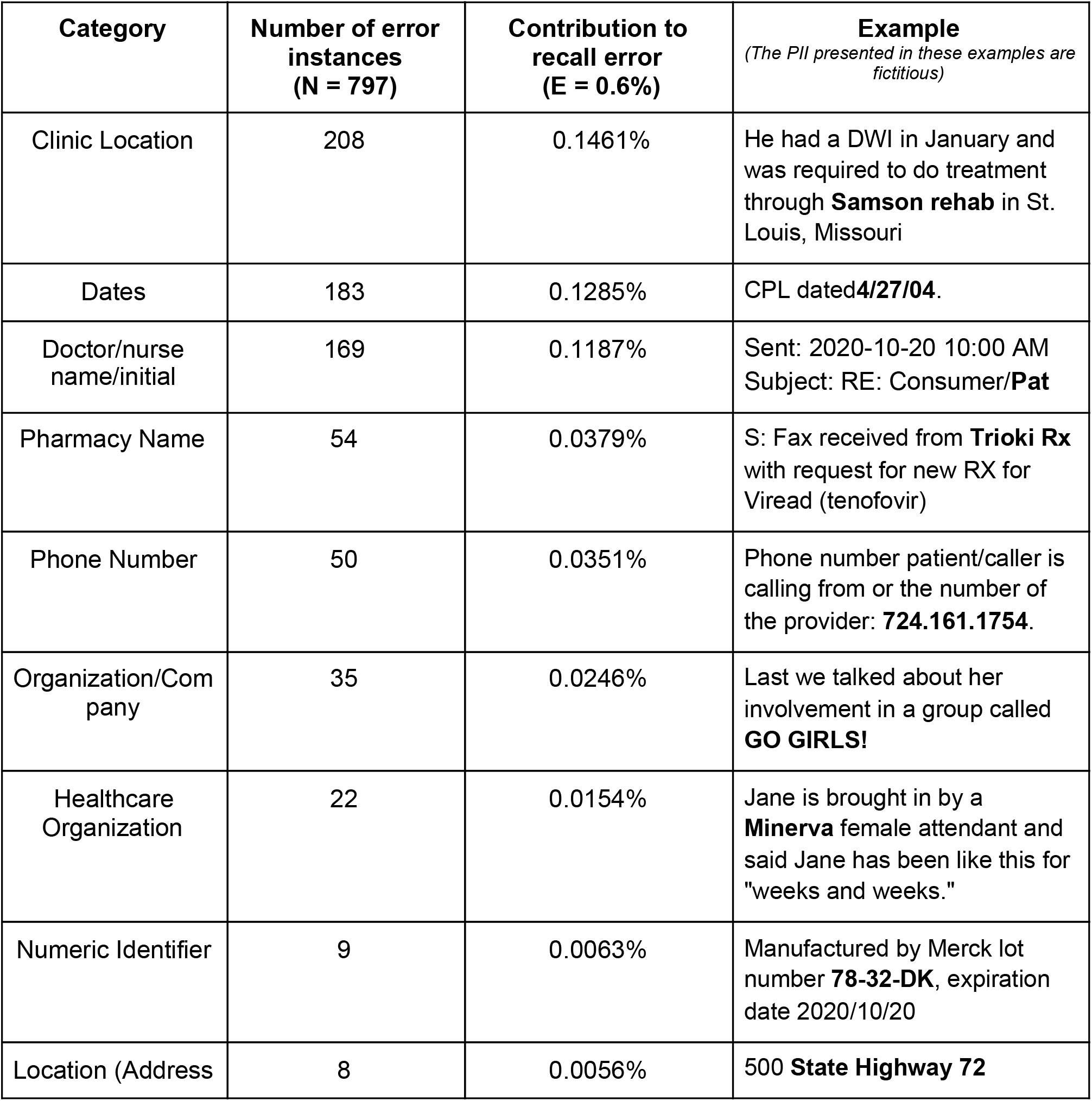
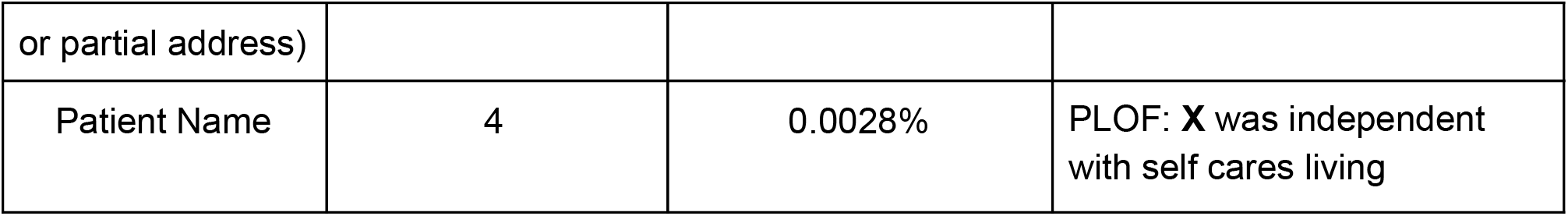
Prevalence and examples of types of false negatives encountered by the nference de-identification system when applied on the Mayo test set. The entity highlighted in bold indicates the word or phrase that the system failed to detect.

The most prevalent error was in the recognition of entities pertaining to clinic locations (208 out of 797). Many of these were due to partially identified phrases (e.g., “*Room 7A*” was missed in “Out of *Southwest Building Room 7A”*). The second most prevalent error type was in dates with 183 false negatives. The third most prevalent error category was in doctor/nurse names and initials with 169 false negatives. Abbreviations and shorthand used by providers (typically while signing off on a clinical note) contributed to the errors in this category

Ambiguous instances of PII also resulted in false negatives. These were cases that a human reader would have difficulty/uncertainty in deeming as PII. An example of this is the word *tp* in the phrase “*Comment: 03-12-2005 08:04:12 - verified tp*”. We found that 26% of errors were those in which the nurse abstractors themselves did not agree on the characterization of PII (Cohen’s Kappa for errors was lower than non-errors, at 0.7453), pointing to the inherent ambiguity.

### Distribution of errors per note

We further investigated the rate at which errors in detecting PII(false negatives) occurred on a per note level. As shown in **Table 5**, the error instances were distributed across 637 notes. Furthermore, we see that a majority of false negatives are spread evenly across the notes (525 out of 637 notes, or 82.4%, contain a single error). For each subsequent error rate, we computed the coverage of PII entities. Here, coverage represents the fraction of PII present in the subset of notes up to the corresponding error rate.

**Table 5:** Distribution of number of errors per note. PII coverage represents the fraction of PII present in the subset of notes up to the corresponding error rate. Average number of error types denotes the number of distinct errors types (such as date errors or name errors) per note.

| Errors per Note | Number of Notes | Total Errors | Cumulative Errors | PII Coverage | Average Number of Error Types |
| --- | --- | --- | --- | --- | --- |
| 0 | 9363 | 0 | 0 | 0.9940 | 0 |
| 1 | 525 | 525 | 525 | 0.9978 | 1.00 |
| 2 | 80 | 160 | 685 | 0.9989 | 1.56 |
| 3 | 10 | 30 | 715 | 0.9991 | 1.75 |
| 4 | 6 | 24 | 739 | 0.9992 | 2.30 |
| 5 | 6 | 30 | 769 | 0.9994 | 2.75 |
| 6 | 2 | 12 | 781 | 0.9995 | 2.5 |
| 7 | 2 | 14 | 795 | 0.9996 | 2.25 |
| 8 | 2 | 16 | 811 | 0.9997 | 2.25 |
| 9 | 3 | 27 | 838 | 0.9999 | 2.33 |
| 10 | 1 | 10 | 848 | 1.0000 | 2.00 |

Even for notes with a large number of errors (more than six), the number of distinct error types is between two and three. This illustrates that most of the errors are of the same type and an artifact of repetition of text within a note. For example, in the note with ten errors, eight of the instances were related to location while the remaining two are related to date. Examples of the errors pertaining to location here are “*Location of INR sample* : *Other: Smallville Other: Smallville Other: Smallville*”, “*Recommend Recheck* : *Other: 04/01/2017 Smallville Other: 04/01/2017 Smallville”, “Recommend Recheck* : *Other: 04/01/2017 Smallville Other: 04/01/2017 Smallville Other: 04/01/2017 Other: 04/01/2017 Smallville”*. Here, the set of location errors all pertain to the same location “Smallville”, which illustrates how the effective amount of identifiable content is substantially smaller than suggested by the raw count. The date presented (“04/01/2017”) was successfully detected. Both the date and location have been replaced with synthetic values for the purpose of this example.

### Distribution of note types

In the Mayo test set, a physician note is associated with a note type (e.g. progress note, emergency visit, telephone encounter). Given that the structure and semantics of these note types vary greatly from each other we analyze the enrichment of errors across them. From the 637 notes with errors, we found 134 distinct note types with at least 1 error. The top 14 note types with highest error content are listed in **Table 6**. Notes of the type “Anti Coag Service Visit Summary” contain the highest rate of errors (22 out of 26 sampled notes) followed by “Electrocardiogram” (19 out of 30 sampled notes).

**Table 6:** Distribution of number of errors per note type. The proportion of sampled notes for a given type that contain at least one error is presented in the last column. This indicates in which note type an error is more likely to occur.

| Note Type | Number of Error Instances | Number of PII Instances | Number of Notes with at Least One Error | Total Number of Notes | Fraction of Notes With at Least One Error |
| --- | --- | --- | --- | --- | --- |
| Phone Message/Call | 60 | 7,466 | 54 | 605 | 0.09 |
| Ambulatory Patient Summary | 59 | 14,502 | 49 | 334 | 0.15 |
| Physician Office/Clinic Message | 42 | 8,352 | 36 | 661 | 0.05 |
| Report | 50 | 3,173 | 36 | 131 | 0.27 |
| Medication Renewal/Refill | 36 | 4,626 | 31 | 358 | 0.09 |
| Progress Note, Family Practice | 27 | 4,975 | 24 | 237 | 0.10 |
| Ambulatory Discharge Medication List | 27 | 8,109 | 23 | 226 | 0.10 |
| Anti Coag Service Visit Summary | 24 | 1,189 | 22 | 26 | 0.85 |
| Electrocardiogram | 19 | 411 | 19 | 30 | 0.63 |
| Anticoagulation Patient Intake - Text | 49 | 5,777 | 18 | 50 | 0.36 |
| Letter | 15 | 3,519 | 14 | 157 | 0.09 |
| Ambulatory Depart Summary | 12 | 3,938 | 12 | 163 | 0.07 |
| Progress Notes | 14 | 3,943 | 11 | 199 | 0.06 |
| Telephone Encounter | 12 | 2,034 | 11 | 273 | 0.04 |

## Methods

### Usage of Mayo Clinic Dataset

The Mayo EHR dataset is based on data from 477,000 patients that originated from multiple EHR data systems (including Epic and Cerner) spanning over 20 years. The dataset includes 104 million physician notes that capture the healthcare journey of patients in addition to structured tables containing lab test measurements, diagnosis information, orders, and medicine administration records. This research was conducted with approval from the Mayo Clinic Institutional Review Board.

We randomly sampled 10,000 notes, which were reduced to the set of unique sentences. This yielded a test set of 172,102 sentences. These were subsequently annotated by six Mayo Clinic nurse abstractors to create a ground truth label for every word and/or phrase. Each sentence was annotated by at least two different nurse abstractors. The inter-annotator agreement on labelling a token as PII had a Cohen’s Kappa of 0.9694 (see Supplementary Methods for details).

An additional set of 10,000 notes were selected to fine-tune the models. We manually annotated 61,800 unique sentences from these notes to create a tagged fine-tuning set. See Supplementary Methods for more details.

### Detection of PII entities

The ensemble architecture described in this section leverages state of the art attention-based deep learning models in conjunction with rules harvested from the data (each of which is described below) to handle semi-structured text. **(Fig. 2)**

**Fig. 2.**
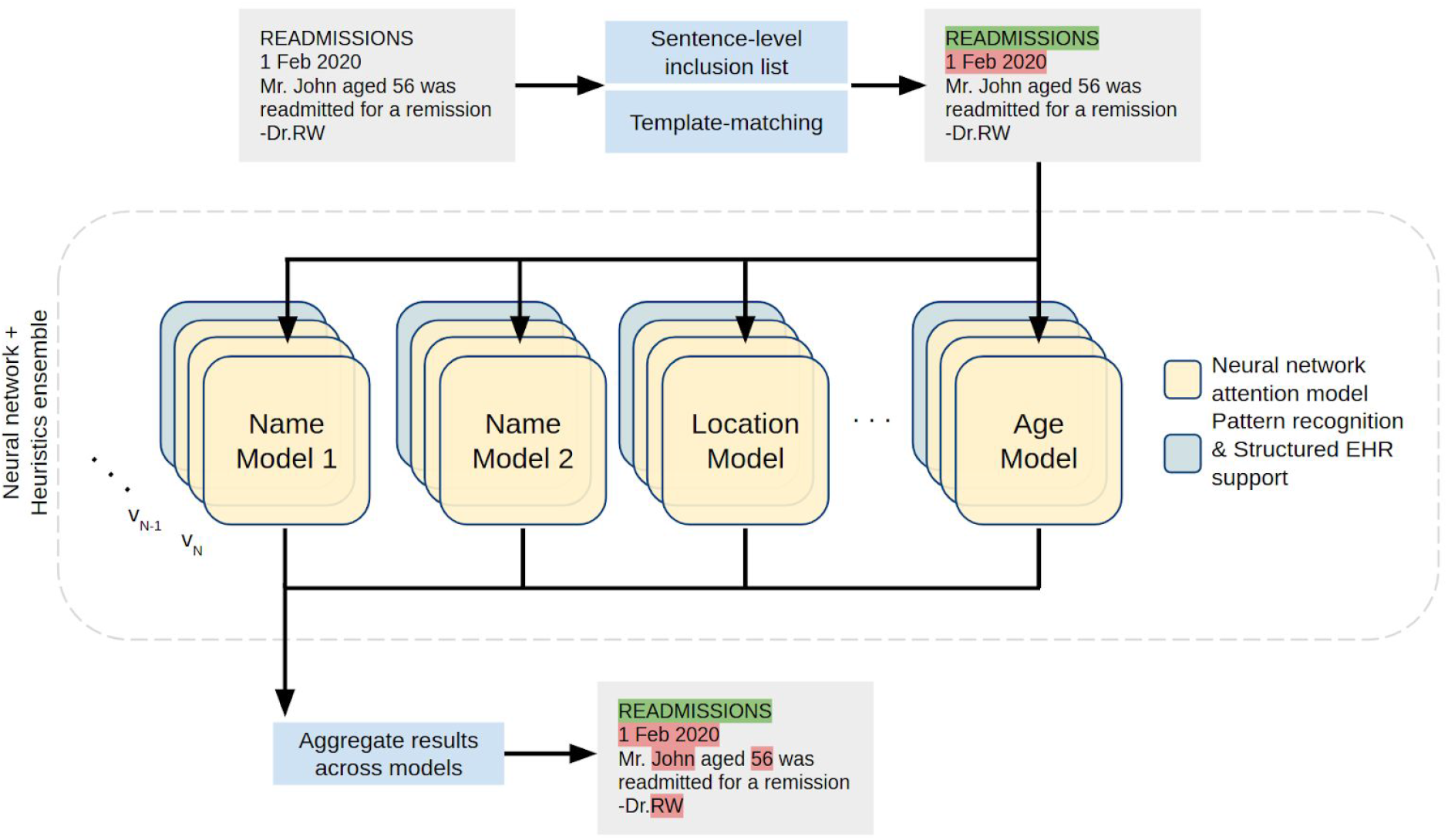
Sentence-based inclusion lists and template matching prune out sentences that either 1) lack PII or 2) contain PII in specific well-defined patterns. An ensemble of attention-based neural networks identify complementary features across different PII types. For each entity type, multiple model versions (*v*_*1*_, *v*_*2*_, *v*_*N*_) are used in tandem. Additionally, pattern recognition modules and structured EHR content from matched patients support the anonymization process. The results from each component of the ensemble are aggregated to yield the original note labelled with PII tags.

There are several salient features of this approach that are worth noting.

### Hybrid Deep Learning Models

The newer breed of attention based deep learning models, in conjunction with transfer learning, allow for faster tuning of these models with significantly smaller sets of labeled data for detecting PII identifiers. We use pre-trained language models based on the BERT^16^ architecture that are then fine tuned for detecting (a) personal names, (b) organizations, (c) locations, and (d) ages. We employed the bert-base-cased model (https://huggingface.co/bert-base-cased) through the HuggingFace/Transformers (https://github.com/huggingface/transformers) library. This is a case-sensitive English language pre-trained model based off of the BERT architecture trained using a masked language modelling (MLM) objective. The fine-tuning process involves training the pre-trained language model on a named entity recognition task using a training set of annotated sentences. We used a total of 61,800 tagged example sentences to fine-tune the models. We fine-tuned each transformer model with a maximum sequence length of 256 (after tokenization) over 4 epochs. We use a training batch size of 32 and a learning rate of 5e-5 with a warmup proportion of 0.4. We then evaluated the model on a validation dataset and computed the accuracy. We performed the fine-tuning and model validation processes in an iterative manner (see Supplementary Methods and **Supplementary Table 1** for complete implementation details). Identifiers such as names, locations, organizations and ages are well suited to a statistical entity recognition method because they can use the context of the surrounding text to disambiguate the entity type of a word. By contrast, pattern matching rules are significantly hampered in this respect. It would be hard, for instance, to detect “Glasgow” as a medical term in “He had no helmet and his Glasgow Score was 6” and as a location in “Mr. Smith had visited his family in Glasgow using lookup dictionaries.

However, we use patterns to deterministically tag reasonably well-defined PII identifiers, which are almost entirely context independent and unambiguous. This category includes dates and times, phone and pager numbers, clinical IDs and numeric identifiers, email, URLs, IP addresses, and vehicle numbers. In addition, harvested sentence templates (described further below) are relied upon to deterministically tag PII instances matched by the template patterns. Our methods apply to content in both structured (e.g lab comments) and free form text (e.g progress notes).

Additionally, it should be noted that we designed our method to detect and transform information about those who provide care, such as physicians, nurses, and pharmacies. Though this is not required by HIPAA Safe Harbor, it allows healthcare organizations to protect the identities of their employees as well.

### Ensemble of models framework and iterative fine tuning

Given the regulatory necessity of extremely high recall for de-identification, we aggregate the results of multiple models trained for the same PII type. Our ensemble involved employing at least one individual model for names, organizations, locations and ages (see **Supplementary Table 2**). An additional text normalized model was also trained and utilized for names. In this respect, if a term is detected as PII in any of the models for that type, then it is tagged. A divide and conquer approach has been implemented that harnesses the power of multiple models to identify PII or extract meaningful entities (***Fig. 2***). In contrast to a “one size fits all” model, this approach enables each individual model to be fine-tuned to learn different (and complementary) features of the unstructured EHR data as has been shown to be used in prior de-identification systems^23^. For instance, one model focuses on identifying peoples’ names while another is geared towards addresses and locations.

Furthermore, there are additional models corresponding to cased and uncased variants of the raw data (referred to as “*Name Model 1*” and “*Name Model 2*” in **Fig. 2**). Each model here corresponds to an attention-based deep neural network. One advantage of carving out the entity space to be handled individually by separate models is that each model needs to only learn the distribution of entities of a specific type as opposed to all entities. However, this introduces a challenge in resolving terms in a sentence that have conflicting and/or ambiguous entity types. These conflicts are resolved in the aggregation phase of our ensemble where a simple voting threshold of one claim is employed (i.e., an entity is considered PII even if one model in the system tags it as such). Since the majority of the components in the ensemble are designed to detect complementary features, we are able to improve recall without much loss of precision.

### Integrating databases as part of core model

We use publicly available databases of names, locations, and addresses to supplement the model fine-tuning process. First names with supporting gender information were downloaded from the US Census database. Cities across the US as well as lists of hospitals were obtained from Wikipedia. These public databases were used to augment training of our models.In addition, patient-specific information from structured EHRs, including patient names and residential addresses, are used to augment the model training and match against PII in the text.

### Sentence-based inclusion list

Clinical note corpora contain a large number of repeated sentences. These stem from various processes, including automated reminders (e.g., “*Please let your doctor know if you have problems taking your medications*”), repeated phrases in the writing style of physicians (e.g. “*Rubella: Yes*”, “*Pain symptoms: No*”) or shared elements in the clinical notes such as section headers (e.g. “*History of Present Illness*”). From the corpus of physician notes from the Mayo Clinic, a set of 1,600 sentences, that did not contain PII, were incorporated into an “inclusion list”. This inclusion list was further expanded with a set of 25,000 sentences containing medically relevant entities, such as disease or drug names (see Supplementary Methods for details on how the inclusion list was constructed). This has the added benefit of improving the precision of the de-identification system because it reduces the risk of misclassifying these important entities as PII by the neural network models. Additionally, sentences marked as being devoid of PII during the validation phase in the iterative fine-tuning process are also added to the inclusion list (see Supplementary Methods).

### Auto-Generating templates using statistical NER models

In addition to exact sentences with high prevalence there are also a large number of PII containing sentences that can be mapped to a template (e.g., *“Electronically signed by: SMITH, JOHN C on 01/02/1980 at 12:12 PM CST”* maps to a template of the form “*Electronically signed by: <LAST NAME>, <FIRST NAME> <INITIAL> on <DATE> at <TIME>“*). While machine learning NER models can be trained and/or fine tuned to learn these patterns, there are instances where entity recognition fails. So, though a name of the form “SMITH, JOHN C” might be detected, “DEWEY” in “DEWEY, JONES K” may not be detected. By contrast, regular expression rules faithfully match every PII for these cases.

The problem, however, is that the process of identifying such templates and generating the corresponding regular expressions is an arduous task because it involves manual inspection of a sufficiently large sample of sentences in the corpus. Here, we use the NER ensemble models designed for the detection of PII to aid in the harvesting of these pattern templates. Sentences from our fine-tuning set of 10,000 notes are passed through the ensemble and detected PII is transformed to its corresponding IOB2 (Inside-outside-beginning) mask (e.g., *“Electronically signed by: B-PER I-PER I-PER on B-DATE at B-TIME PM CST”*) generating a potential NER template. Additionally, a ‘syntax template’ for these sentences is also generated, such that any term that was detected as an entity is mapped to its syntactic representation - one of ‘W’ for alphabets only, ‘N’ for numbers only and ‘A’ for alphanumeric (e.g., *“Electronically signed by: W, W W on N/N/N at N:N PM CST”*). Finally, for each unique syntax template, if there exists only one NER template amongst all instances of the syntax template, a regular expression rule is generated (e.g *“Electronically signed by: [A-Za-z]+, [A-Za-z]+ [A-Za-z]+ on \d+/\d+/\d+ at \d+:\d+ PM CST”*) by mapping each syntax token to its corresponding regular expression pattern - ‘W’ to ‘[A-Za-z]+’, ‘N’ to ‘\d+’ and ‘A’ to ‘\w’.

### Transformation of tagged PII entities

The de-identification process is designed to recognize words and phrases that represent PII and other sensitive elements with high recall. However, if the input text is transformed to the de-identified version by *redacting* detected PII, undetected PII (e.g., ‘Hayley’ and the date ‘7/21’ in **Fig. 3**) is obviously leaked to any person who reads the document. As such, the obfuscation process aims to conceal these residual PII by *replacing* detected PII with suitable surrogates so it is difficult to distinguish between the residual PII and the surrogates^21,24,25^. This method has been implemented in several de-identification approaches^26,27^. As highlighted in **Fig. 3**, it is difficult for a human to determine which of “Jack Michaels” or “Hayley” is a leaked instance of PII in the output of the replacement strategy using this mechanism of Hiding in Plain Sight (HIPS)^28^. Evidence with human readers has shown that when the recall of a natural language processing tool is high (i.e., when most real identifiers are detected), the rate of distinguishing real from filler identifiers is no better than what one would encounter by random chance. It has further been shown, however, that under highly controlled conditions, it is possible for a machine learning system to replicate the behavior of the natural language de-identification tool to remove fillers and leave real identifiers in place^28,29^.

**Fig. 3.**
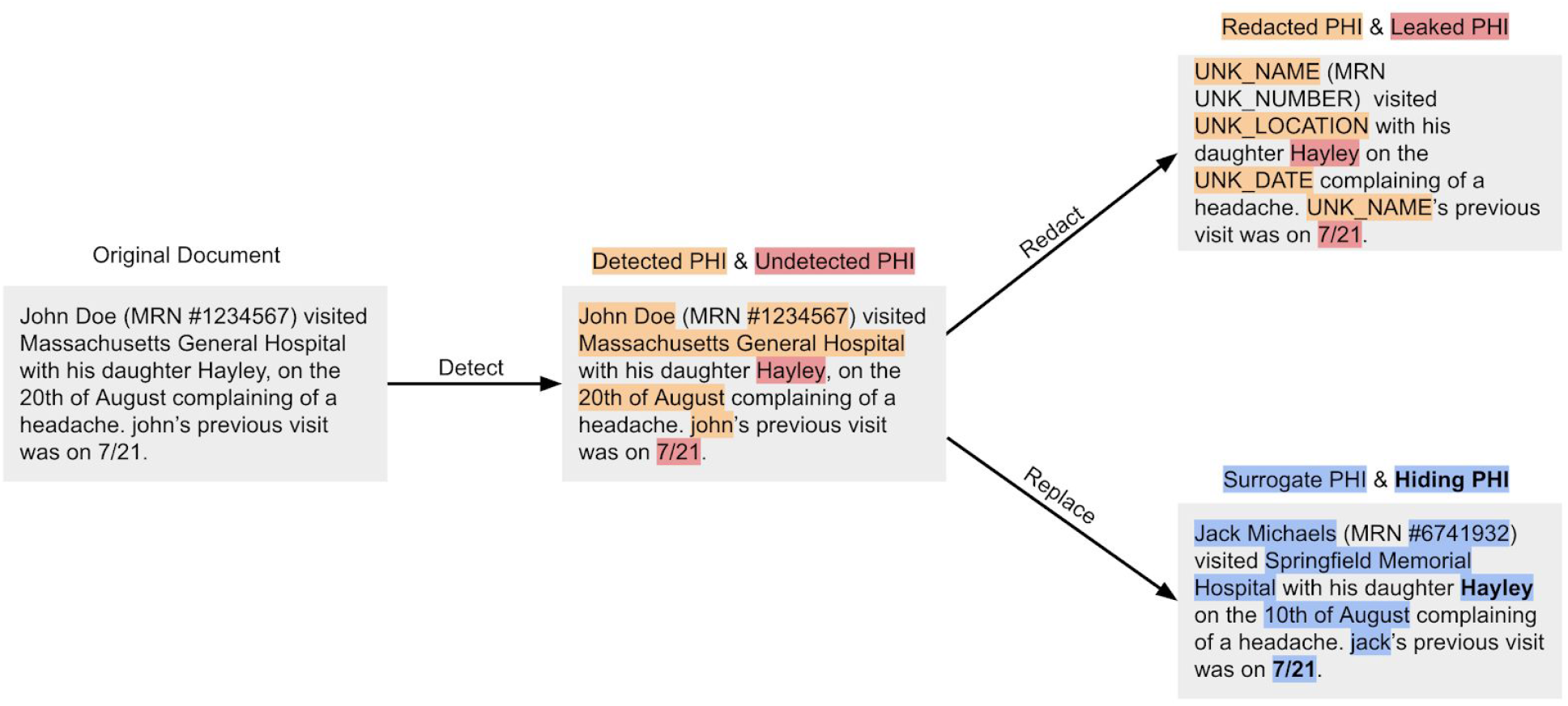
An illustration of the hiding in plain sight (HIPS) mechanism to highlight the utility of the detect → replace strategy. After obfuscation, distinguishing real PII from surrogates is no better than what one would expect by random chance.

In addition to employing the HIPS method, we apply entity-specific rules and heuristics to improve the fidelity of the surrogate. We further improve interpretability of the output by ensuring that every unique PII token in all EHR records for a patient has the same transformation. Consider the input text “*John Smith, a pleasant 67 year old presented with his son Jack. John complains of breathing difficulty*” was transformed to “*Jane Kate, a pleasant 67 year old presented with his son Matt. Ryan complains of breathing difficulty*.” In this example, “*Jane Kate*” as a surrogate is an obvious giveaway that it is a fake name and therefore lends itself to be distinguished from any true PII that may have leaked. Furthermore, it appears that a third completely different person is complaining of breathing difficulty. So an ideal transformation would have maintained the format of first name followed by last name and the gender for “John Smith” and every instance of “*John*” or “*Smith*” in the input would be transformed to the same output; something like “*Jacob Hamilton, a pleasant 67 year old presented with his son David. Jacob complains of breathing difficulty*.”

As discussed, we manage the replacement of surrogates per entity type (see Supplementary Table 2). Names are transformed in a manner that is consistent with format, gender and ethnicity of the original (i.e., “*Ms. Lopez visited New York General Hospital for her routine checkup”* becomes “*Ms. Hernandez visited Mass General Hospital for her routine checkup”*). Dates are handled in a way to preserve their formatting (i.e., “*March 5th, 2014*” becomes “*February 27th, 2014*” and “*03-05-2014*” becomes “*02-27-2014*”). The shift in the date is a patient-specific random number. This ensures that dates are shifted consistently for a given patient. Locations and organizations are replaced with suitable surrogates chosen from a predefined dictionary. PII entities that contain numeric digits (such as phone number or patient ID) involve replacing these numbers randomly while maintaining overall length and format.

While the transformation output of an input token is the same for all instances of its occurrence for a given patient, they would be different across patients. That is, while all instances of “John” in one patient might be transformed to “Jacob” for another patient it could be “Aaron”.

## Discussion and Future Work

Numerous approaches to de-identification have been developed. Automated de-identification systems can broadly be segmented into four categories: (i) rule-based systems, (ii) traditional machine learning systems, (iii) deep learning systems and (iv) hybrid and ensemble systems.

**Rule-based systems**^19,20,30–32^ use pattern matching rules, regular expressions, dictionary and public database lookups to identify PII elements. These are simple to implement and usually deterministic; however, these systems have several drawbacks. First, pattern matching rules for identifiers are typically not robust for handling variance in input due to typographical errors(spelling, punctuation, casing etc.); A rule that matches *“Dr. John”* may not be able to match *“Dr john”*. Second, creating template patterns to match sentence fragments like *“Provider Name: Dr. John”* that tag any term after *“Provider Name: Dr*.*”* as a name, for example, requires manual effort to understand the data to create these templates. Doing this for large data sets with notes for millions of patients is time consuming and intractable. Third, dictionary-based systems may not be complete, resulting in increased ‘false negatives’ (i.e. true PII that is not detected). Fourth, blindly using dictionary/database lookups induces ‘false positives’ because they tag phrases that are not identifiers in the context in which they are used that need to be disambiguated^33^. For example, in *“The doctor determined his Braden Score as normal”*, the term “Braden” might be flagged as PII, when it is only a clinical term.

**Traditional Machine Learning (ML) systems**^21,34–36^use traditional machine learning (ML) algorithms, such as support vector machines (SVMs) and conditional random fields (CRFs), to perform NER classification as PII for each word in a sentence. The classification task involves creating labeled data and defining features based on properties like part of speech (POS) tags, typography (e.g., capitalization, casing, spacing, font weights, or font types), punctuation, and frequency of words and/or their neighbors. These methods, in addition to requiring significant effort in encoding the feature vectors, may not generalize across datasets.

**Deep Learning systems**^18^, have become the state-of-the-art for a wide variety of application domains, including vision (e.g., image classification) and speech (e.g., voice recognition and generation). In language-related tasks (e.g machine translation), these approaches have surpassed human level performance^37^. Deep learning has proven beneficial in numerous NLP tasks, including predicting the next word (language modeling), tagging tasks such as part of speech tags, entities in a sentence (entity recognition), and dependency parsing. This has enabled applications that traditionally required custom rules and hand-crafted features to be solved without any feature engineering. Modern deep learning approaches for de-identification have been shown to outperform their predecessors^18^, but they require very large quantities of domain specific labeled training data to perform well. Specifically, the challenges include, but are not limited to, the presence of long and highly descriptive sentences, usage of clinical shorthand (that vary across physicians and medical specialties), and a variety of semi-structured machine generated content. Moreover, publicly available datasets for de-identification (including the popular i2b2 2014 dataset^17^) lack diversity, often focusing on only a few types of notes or areas of disease. Training and benchmarking with such datasets is likely to bias the resulting models and fail to capture the nuanced and complex nature of physician notes. Recently, attention-based neural network (transformer) models have also been implemented for de-identification but have shown limited generalizability in the absence of support from encoded rules^38^.

**Hybrid^39^ and Ensemble Systems**^40,41^ use combinations of rule-based and machine learning-based components in tandem to improve PII detection efficacy. With these approaches, the choice of components, finding the right split of tasks between them and the optimal strategy for combining results from them become crucial. Some approaches^42^ invoke engineering post-processing layers that fix the errors that are introduced by other (earlier) components. In cases where there is, by design, overlap in the type of PII being predicted (e.g. multiple components detecting people names), considerable effort is spent measuring and choosing a method, like a stacked meta classifier or voting scheme, to pick a winning component^40^. The nference de-identification system presented here addresses the limitations of prior methods^11^ and achieves high levels of recall and precision.

There are several opportunities to further improve the performance of de-identification systems. First, existing knowledge graphs and language models trained on biomedical corpora can be leveraged. For example, if a patient’s note contains the sentences “*Patient diagnosed with lung cancer” and “ECOG performance status was determined to be 2*”, ECOG would not be treated as PII since it has a strong biological association with lung cancer based on the knowledge graph. In the de-identification process, this could be used to recover biological terms incorrectly tagged as PII (false positives). Second, the quality of sentences that are provided to the model can be improved. Unstructured clinical text does not always contain well-formatted text commonly due to missing punctuations and incorrect casing. A case-sensitive pre-trained model along with a masked-language model objective can be used to train a system capable of correctly introducing punctuation in the right location. Another challenge with the quality of clinical documents is the prevalence of short fragments and bullet points giving rise to sentences with poor context. Context of a single sentence can be expanded using preceding and succeeding sentences or employing document level transformer models such as Transformer-XL^43^. Third, unsupervised methods can be incorporated to accelerate the annotation process of the NER task. Grouping the word representations generated by a transformer model yields informative clusters (e.g. a cluster of names) that can be annotated according to the nature of words present in the cluster. The NER task can then be formulated as a mask language task, where the overlap of the list of potential candidates for a missing word with the clusters can inform the entity type of the missing word.

## Conclusion

Overall, this work implemented an ensemble approach to de-identification of unstructured EHR data incorporating transformer models supported by heuristics for automatically identifying PII across diverse clinical note types. Upon detection, suitable surrogates replaced PII in the processed text thereby concealing residual identifiers (hiding in plain sight). The system demonstrates high precision and recall on both publicly available datasets and a large and diverse dataset from the Mayo Clinic.

## Data Availability

2014 I2B2 data is publicly available subject to signed safe-usage and research only.
The Mayo EHR clinical notes are not publicly available at this time.

## Acknowledgements

We would like to thank the Mayo Clinic and the Mayo Clinic IRB under whose auspices the development of the de-identification methods and testing against real world datasets was made possible. We thank the nurse abstractors - Wendy Gay, Kathy Richmond, Denise Herman, and Sandra Severson, Dawn Pereda and Jane Emerson - for annotating the ground truth for the 172, 102 sentences in the Mayo dataset that was used for testing the performance of the system, the Mayo Data Team of Ahmed Hadad, Connie Nehls and Salena Tong for preparing and helping us understand the Mayo EHR data and Andy Danielsen for supporting the collaboration. Finally, we thank Murali Aravamudan, Rakesh Barve and A. J. Venkatakrishnan for their thoughtful review and feedback on the manuscript.

## Disclosures

Jeff R. Anderson, John D. Halamka, and William A. Faubion Jr. do not have any conflicts of interest in this project. Bradley Malin is a contracted consultant of the Mayo Clinic. Karthik Murugadoss, Ajit Rajasekharan, Vineet Agarwal, Sairam Bade, Jason L. Ross, Venky Soundararajan, and Sankar Ardhanari are employees of and have a financial interest in nference. Mayo Clinic and nference may stand to gain financially from the successful outcome of the research.

## Supplementary Methods

### Ensemble Architecture Implementation details

We employed the *bert-base-cased* model (https://huggingface.co/bert-base-cased) through the HuggingFace/Transformers (https://github.com/huggingface/transformers) library. This is a case-sensitive English language pre-trained model based off of the BERT architecture trained using a masked language modelling (MLM) objective. The BERT model was pretrained on BookCorpus (https://huggingface.co/datasets/bookcorpus), a dataset comprising 11,038 unpublished books in addition to English Wikipedia.

Our ensemble involved employing at least one individual model for names, organizations, locations and ages. An additional *text normalized* model was also trained and utilized for names. Here, text normalization refers to the process of converting all uppercase words to title case (lowercase words are retained as is). A total of 61,800 tagged example sentences were used for fine-tuning the models. The final number of examples for each entity type is shown in *Supplementary Table 1*.

**Supplementary Table 1:** BERT models employed in our ensemble and the corresponding entity type and number of fine-tuning examples. The Model Priority # denotes the order of precedence in the event that a word is tagged as PII by multiple models. For example, if a word is tagged as both a name and a location, it will be assigned the name entity (which has higher priority).

| Model Priority # | Entity Type | Text Normalized? | Fine-tuning Examples |
| --- | --- | --- | --- |
| 1 | Name | No | 44,929 |
| 2 | Name | Yes | 44,929 |
| 3 | Location | No | 11,461 |
| 4 | Age | No | 5,409 |
| 5 | Organization | No | 44,825 |

Each transformer model is fine-tuned with a maximum sequence length of 256 (after tokenization) over 4 epochs. We use a training batch size of 32 and a learning rate of 5e-5 with a warmup proportion of 0.4. The Adam optimization algorithm was employed to update network weights. Loss was computed using cross entropy loss.

Each model is iteratively fine-tuned with training samples being continuously added to the initial set of training samples. The sentences chosen for fine-tuning the model are specifically selected from the space of errors that was seen in prior models. The iterative process of fine-tuning models therefore results in the generation of multiple individual neural networks (different versions) for each PII type each having a specific performance. To maximize the overall recall, we choose the two best performing models for each entity type and employ them in tandem.

To complement the above improvements on model architecture and algorithms for de-identification, an iterative learning framework is deployed in tandem that allows rapid validation and performance evaluation for trained models (***Supplementary Fig. 1***). This allows each component of the ensemble framework to be re-trained and fine-tuned to learn from previous mistakes independently of other models.

**Supplementary Fig. 1:**
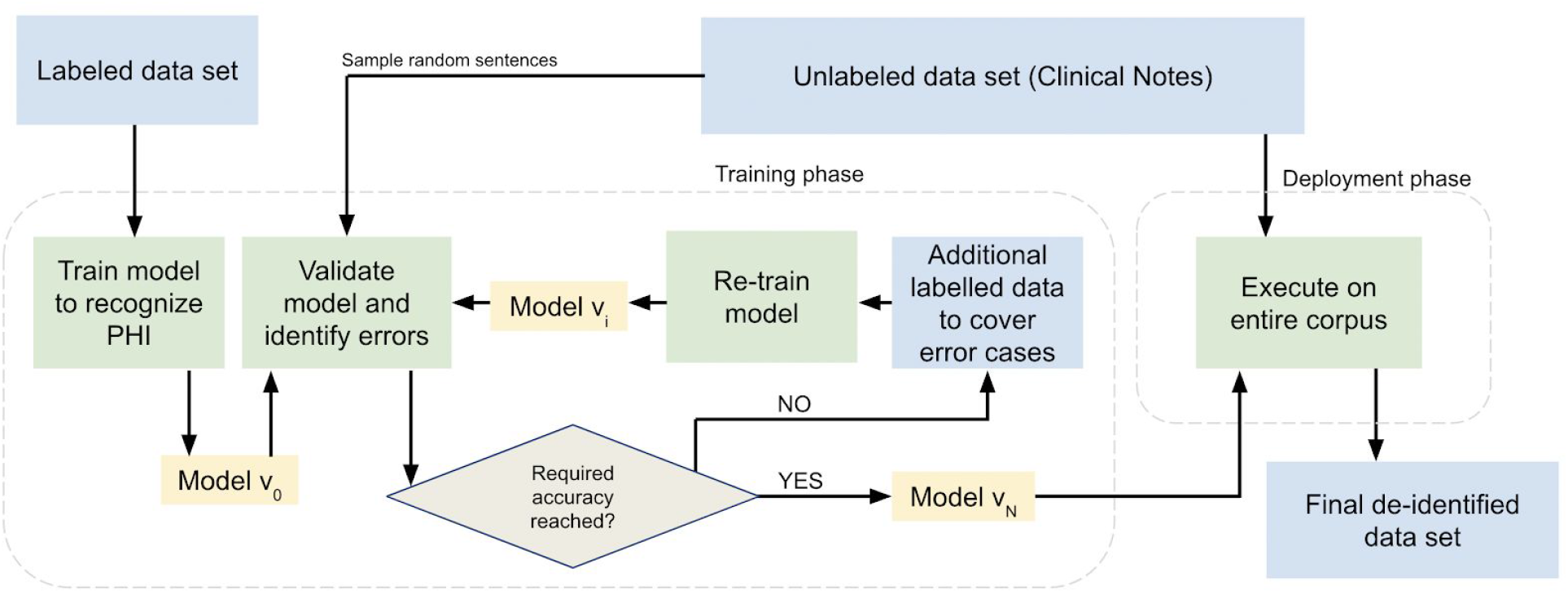
Iterative model generation process and learning from errors. Model performance improves during its evolution from v0 to vN.

All of our experiments were performed on an Ubuntu 16.04 machine (12 CPU cores and 220GB RAM) with two NVIDIA Tesla V100 GPUs (16GB of RAM each). We used Python v3.6.9 with PyTorch v1.3.1 and pytorch-pretrained-bert v0.6.1 (now HuggingFace/Transformers).To de-identify text, we first perform sentence tokenization to convert documents into sentences. On two GPUs, our system achieved an inference speed of 53 sentences per second (inference batch size was set to 128). Additionally, fine-tuning an individual model of our ensemble took 45 minutes for ∼44k sentences with both GPUs being utilized.

In order to maximize recall of our ensemble, we employ a voting ensemble scheme across models of different entity types with a voting threshold of 1. That is, a word is determined to be PII if it is detected by at least one model. If a word is detected as PII by more than one model, it is assigned an entity type based on its priority (as described in ***Supplementary Table 1***).

### Creating an inclusion list of sentences

In a repository of 103 million physician notes (from 477,000 patients) from the Mayo Clinic, a total of approximately 3.1 billion sentences corresponded to approximately 700 million unique sentences, which highlights the redundancy in a corpus of this size and provides optimization opportunities in the de-identification processing pipeline. In particular, sentences with high prevalence were found to typically not contain PII (since they occur across a large number of patients, the chances that they contain information specific to any one patient is low). We computed the prevalence of all sentences and found that the top 1,600 most common sentences correspond to 1.01 billion sentences overall (one-third of the entire corpus).

These 1,600 sentences represented the initial inclusion list. Additionally, we filtered out the top 25,000 most prevalent sentences that contain a disease or a drug entity. This ensures that medically relevant sentences that are also highly prevalent are preserved. All of the sentences that are part of the inclusion list are manually verified.

### Obfuscation methods

For each category of PII, obfuscation is performed through the replacement methods described in Supplementary Table 2.

**Supplementary Table 2:**
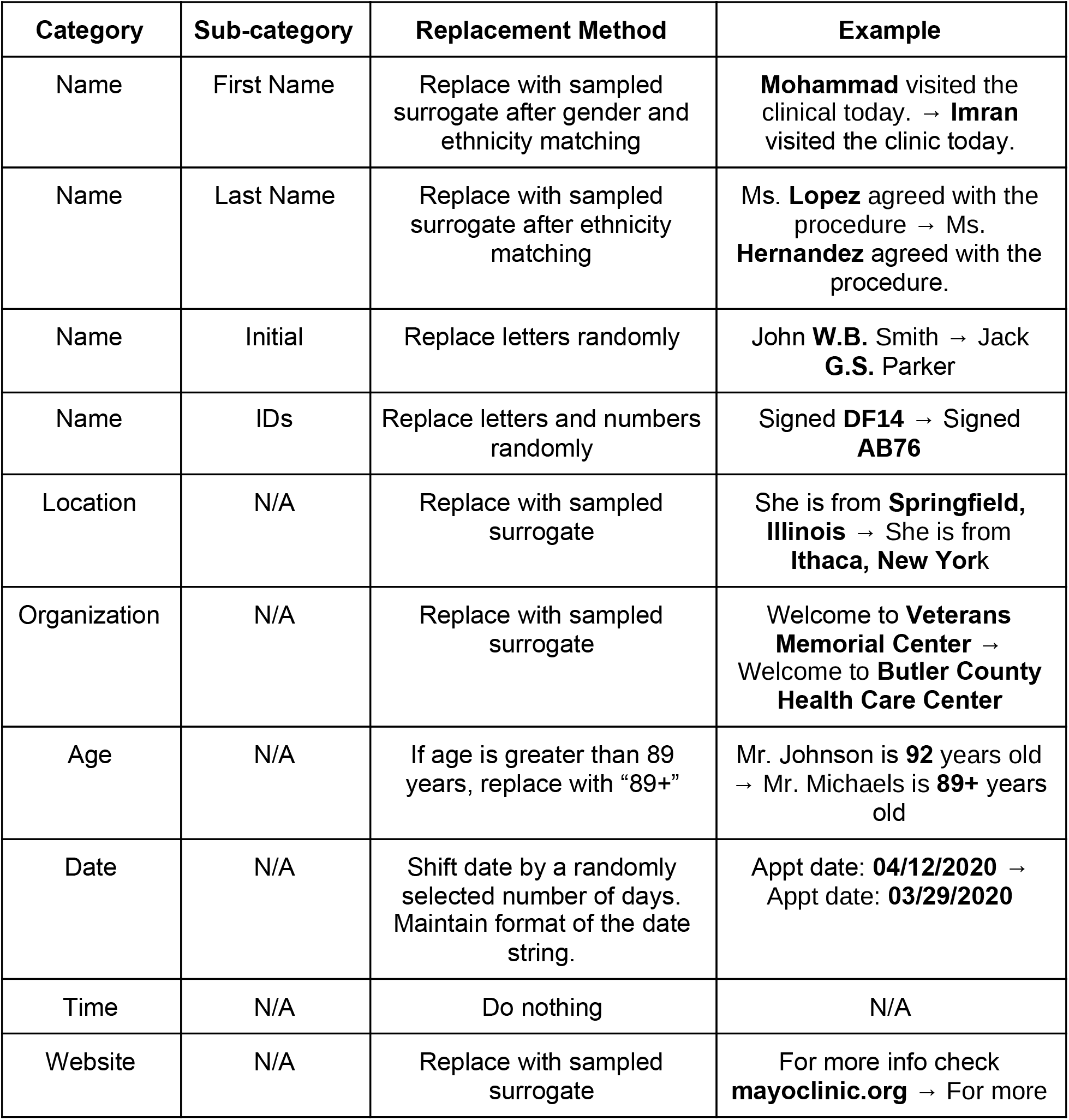
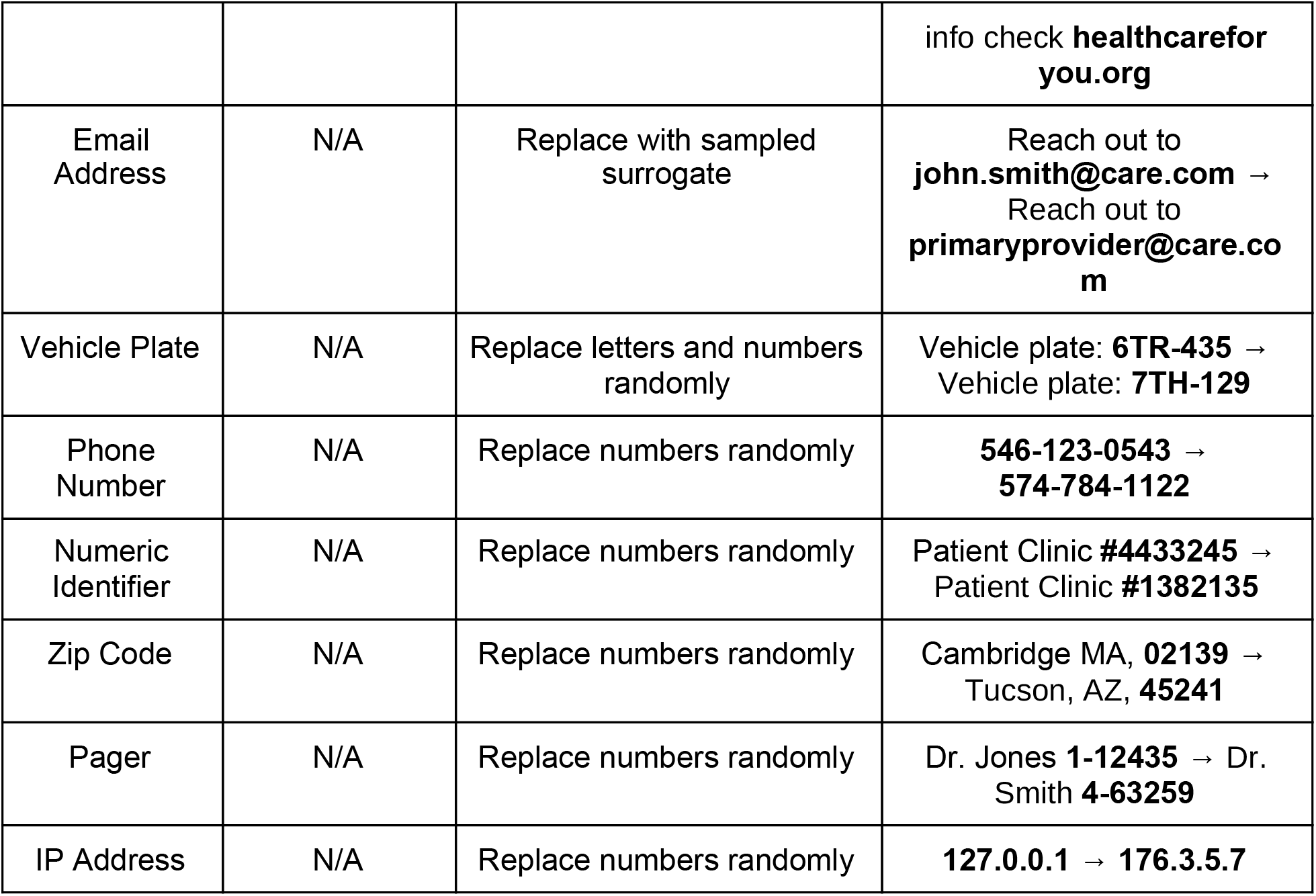
Obfuscation methods for each PII category

### Evaluation metrics

To evaluate model performance on the de-identification task, we computed the precision, recall and F1 scores. These were computed as follows:

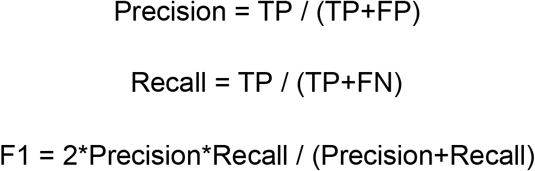

where TP is the true positive count, FP is the false positive count and FN is the false negative count.

### De-identification on 2014 I2B2 test dataset

The 2014 I2B2 dataset consisted of 515 notes each in an individual XML file (present in the folder:. /2014 De-identification and Heart Disease Risk Factors Challenge Downloads/test_data/PHI Gold Set - Fixed).

### Evaluation of existing methods

We report the performance of Scrubber, Physionet and Philter systems on the 2014 I2B2 data in their standard modes of operation (without additional dictionaries or gazetteers). To run MIST on the 2014 I2B2 data, we converted the dataset into the 2006 I2B2 data format since the stable software release of MIST directly supported the 2006 format (and not the 2014 format). Additionally, MIST assigns PII categories that are different from the 2014 I2B2 entity set. To address this issue, we constructed a mapping between the two sets of PII categories as described in **Supplementary Table 3**. In our implementation of MIST, we did not use gazetteers. As a result the scores we report for MIST are lower than those of the Dernoncourt et al. implementation which was configured to use the same gazetteers as their CRF model. We installed and implemented NeuroNER with instructions as outlined in the GitHub repository (https://github.com/Franck-Dernoncourt/NeuroNER/). In particular, we downloaded and ran the *i2b2_2014_glove_spacy_bioes* pre-trained model on the I2B2 validation set.

**Supplementary Table 3:** Mapping between MIST and I2B2 PII categories

| MIST PII Category | I2B2 PII Categories |
| --- | --- |
| NAME | PATIENT, DOCTOR, USERNAME |
| LOCATION | ORGANIZATION, STREET, CITY, STATE, COUNTRY, ZIP, LOCATION-OTHER |
| AGE | AGE |
| DATE | DATE |
| CONTACT | PHONE, FAX, EMAIL |
| ID | IDNUM, MEDICALRECORD, DEVICE |
| PROFESSION | PROFESSION |

### Handling document IDs

The nference system was designed to identify document IDs in unstructured text (e.g. “*3-1272852*” in the sentence “*eScription document: 3-1272852 BFFocus*”). These entities were however not marked as PII in the ground truth of the I2B2 dataset and hence contributed to the false positive rate of our system. If we exclude such cases (we found 87 instances of document ID) our precision improves from 0.979 to 0.986.

### PII entity-wise precision and recall comparison

For each entity class and I2B2 entity type we computed the precision and recall for both versions of the nference system (fine-tuned only on Mayo data and fine-tuned on Mayo as well as I2B2 data) as shown in Supplementary Table 4. Since the tagset used by nference is different from I2B2 entities, the recall could be calculated for each I2B2 entity and for each entity class. However, the precision could only be determined at the level of the entity class. Rule-based components on the nference ensemble performed identically across both versions of our system since they are not impacted by fine-tuning. Support was computed at the word level (i.e. “John Smith’’ corresponds to a support of 2).

**Supplementary Table 4:** PII entity-wise precision and recall for both versions of the nference system: (a) Fine-tuned on Mayo and (b) Fine-tuned on Mayo+I2B2. The first column corresponds to the entity class and the second column corresponds to the specific I2B2 entity type. Dates, numeric identifiers and contacts are implemented through rule-based methods and therefore have the same precision and recall across both system versions. For this analysis, only ages over 89 in the test dataset were considered (totally 8 instances of such an age were found) and our method detected all of those entities successfully. We therefore omit ages from this table. The tagset used by nference groups is different from I2B2 entities. Therefore, recall is calculated for each I2B2 entity and for each entity class but the precision is determined only at the level of the entity class. (*) While precision and recall have been computed for COUNTRY, STATE and HOSPITAL entities, we do not include for computing the final recall (in accordance with the group B entity set defined in Table 1.)

|  |  |  | nference (fine-tuned on Mayo) |  | nference (fine-tuned on Mayo+I2B2) |  |
| --- | --- | --- | --- | --- | --- | --- |
| Entity Class | I2B2 Entity | Support | Precision (False Positive Count) | Recall (False Negative Count) | Precision (False Positive Count) | Recall (False Negative Count) |
| All | All | 10861 | 0.961 (436) | 0.988 (135) | 0.979 (239) | 0.992 (92) |
| Date |  | 4951 | 0.975 (126) | 0.994 (27) | 0.975 (126) | 0.994 (27) |
|  | DATE | 4951 | N/A | 0.994 (27) | N/A | 0.994 (27) |
| Names |  | 4131 | 0.974 (109) | 0.991 (36) | 0.996 (17) | 0.994 (23) |
|  | PATIENT | 1353 | N/A | 0.992 (11) | N/A | 0.998 (2) |
|  | DOCTOR | 2691 | N/A | 0.992 (21) | N/A | 0.993 (17) |
|  | USERNAME | 87 | N/A | 0.954 (4) | N/A | 0.954 (4) |
| Location |  | 1177 | 0.911 (113) | 0.980 (24) | 0.968 (38) | 0.987 (15) |
|  | STREET | 415 | N/A | 0.978 (9) | N/A | 0.992 (3) |
|  | CITY | 327 | N/A | 0.982 (6) | N/A | 1.0 (0) |
|  | STATE* | 188 | N/A | 1.0 (0) | N/A | 1.0 (0) |
|  | COUNTRY* | 94 | N/A | 1.0 (0) | N/A | 1.0 (0) |
|  | ZIP | 133 | N/A | 1.0 (0) | N/A | 1.0 (0) |
|  | LOCATION-OTHER | 20 | N/A | 0.55 (9) | N/A | 0.6 (12) |
|  |  |  | nference (fine-tuned on Mayo) |  | nference (fine-tuned on Mayo+I2B2) |  |
| Entity Class | I2B2 Entity | Support | Precision (False Positive Count) | Recall (False Negative Count) | Precision (False Positive Count) | Recall (False Negative Count) |
| Organization |  | 1639 | 0.969 (43) | 0.815 (302) | 0.991 (13) | 0.914 (140) |
|  | HOSPITAL* | 1502 | N/A | 0.821 (269) | N/A | 0.922 (128) |
|  | ORGANIZATION | 137 | N/A | 0.759 (33) | N/A | 0.912 (12) |
| Numeric Identifiers |  | 576 | 0.926 (45) | 0.977 (13) | 0.926 (45) | 0.977 (13) |
|  | IDNUM | 201 | N/A | 0.968 (7) | N/A | 0.968 (7) |
|  | DEVICE | 10 | N/A | 0.9 (1) | N/A | 0.9 (1) |
|  | MEDICAL RECORD | 365 | N/A | 0.986 (5) | N/A | 0.986 (5) |
| Contact |  | 171 | 1.0 (0) | 0.988 (2) | 1.0 (0) | 0.988 (2) |
|  | PHONE | 167 | N/A | 0.994 (1) | N/A | 0.994 (1) |
|  | FAX | 3 | N/A | 0.666 (1) | N/A | 0.666 (1) |
|  | EMAIL | 1 | N/A | 1.0 (0) | N/A | 1.0 (0) |

### Mayo test set annotation

#### Inter-rater reliability

Cohen’s Kappa is used to compute the inter-rater reliability for categorical terms. We calculate Cohen’s Kappa for the Mayo test dataset annotated by Mayo Clinic nurses in the following manner.

<u>Step 1</u>: In the ground truth tagged sentences for each nurse, we convert each PII entity (e.g., names, dates, and locations) to a universal “PII entity” type. Non PII entities are left as is.

<u>Step 2</u>: Since the full set of sentences to review is split into three groups and within each group every sentence is reviewed by two nurses, we consider two nurse extractor groups. Group 1 is comprised of nurses 1, 3, and 5 and group 2 is comprised of nurses #2, #4, and #6.

<u>Step 3</u>: We then construct an agreement/disagreement matrix. The numbers in the **Supplementary Table 5** denote the number of words for each category. For example, there are 4,919 words that were marked as PII by group 1 but were not marked as PII by group 2.

**Supplementary Table 5:** Agreement matrix for measuring inter-rater reliability

|  |  | Nurse Extractors Group 2 |  |
| --- | --- | --- | --- |
|  |  | PII entity | Non PII entity |
| Nurse Extractors Group 1 | PII entity | 185455 (a) | 4919 (b) |
|  | Non PII entity | 5411 (c) | 1483221 (d) |

<u>Step 4</u>: The observed proportionate agreement p_0_ = (a+d)/(a+b+c+d) = **0**.**9938**

<u>Step 5</u>: The expected probability (i.e. probability of random agreement between the two groups) is the probability that both groups agreed on either yes or no. The probability that both groups agreed on yes (p_yes_) is given below

Pyes = (a+b)/(a+b+c+d). (a+c)/(a+b+c+d) = **0**.**0128**

P _no_ = (c+d)/(a+b+c+d). (b+d)/(a+b+c+d) = **0**.**7858**

Therefore,

p _e_ = p _yes_ + p _no_ = **0**.**7987**

<u>Step 6</u>: Compute Cohen’s Kappa

κ = (p _o_ - p _e_)/(1 - p _e_) = **0**.**9694**

## Notes

### Funding Statement

No external funding

### Author Declarations

This research was conducted with approval from the Mayo Clinic Institutional Review Board 20-003235 - CDAP: Data De-identification Methods Development. All analysis of EHRs was performed in the privacy-preserving environment secured and controlled by the Mayo Clinic. nference and the Mayo Clinic subscribe to the basic ethical principles underlying the conduct of research involving human subjects as set forth in the Belmont Report and strictly ensures compliance with the Common Rule in the Code of Federal Regulations (45 CFR 46) on Protection of Human Subjects. For more information, please visit https://www.mayo.edu/research/institutional-review-board/overview

